# Poly-Social Risk for Hypertension Among Black and Latina Women

**DOI:** 10.64898/2026.06.12.26355558

**Authors:** Lauren Belak, Kaitlyn James, Lysa Auguste, Marya Rana, Imuetiyan Eweka, Nore De Moor, Amy Sarma, Giulia Ensing, Katherine Economy, Camille Powe, Michael C. Honigberg

## Abstract

**Background:** Hypertension is a leading modifiable cardiovascular risk factor prominently influenced by health-related social needs (HRSN). Whether detailed information on HRSN can improve identification of hypertension among minoritized women is unknown.

**Methods:** Black and Latina women aged 18-65 years completed the Centers for Medicare and Medicaid Services Accountable Health Communities Screening Tool, assessing 13 HRSN domains. Hypertension was ascertained by a validated EHR-based algorithm or self-report of hypertension. Logistic regression tested associations of HRSN with hypertension. LASSO regression with 10-fold cross-validation was used to derive a poly-social risk score in the training set (random 70%) and tested in the validation set (30%) against a sociodemographic model (age, race, income, education).

**Results:** Among 1302 participants (mean [SD] age 40.1 [11.3] years, 70.4% Black, 44.3% Latina), higher cumulative burden of HRSN was associated with increased odds of hypertension (adjusted odds ratio [aOR] for each additional domain of HRSN: 1.07 [95% CI 1.01-1.14], P=0.02). Food insecurity (aOR 2.30 [1.37-3.87], P= 0.002), lapse in utilities (aOR 1.44 [1.04-1.96], P=0.02), poor concentration (aOR 1.57 [1.13-2.17], P=0.007), and social isolation (aOR 1.77 [1.14-2.73], P=0.01) were associated with hypertension. In the validation set, the poly-social risk score did not improve discrimination for hypertension vs. the sociodemographic model (AUC 0.76 [95% CI 0.71-0.81] vs. AUC 0.80 [0.75-0.85]).

**Conclusions:** In this cross-sectional analysis of Black and Latina women, greater cumulative social disadvantage was associated with hypertension. While inclusion of HRSN did not improve hypertension prediction beyond conventional sociodemographic indices, findings may inform targeted interventions among minorities at cardiometabolic risk.

## INTRODUCTION

Cardiovascular disease (CVD) is the most common cause of death among women in the United States, and hypertension is the leading modifiable risk factor for morbidity and mortality prevention^1–3^. Hypertension results from the combined effects of genetics and environment^1,4^, although social factors have become increasingly recognized as strong predictors of cardiovascular outcomes^5–7^.

Despite its prevalence among nearly half of the American adult population^8^, hypertension disproportionately affects racial/ethnic minorities, including 55.3% of Black women and 40.8% of Hispanic/Latina women^1^. Black women are also diagnosed with hypertension at younger ages and have increased risk for hypertension-related complications, including stroke, heart failure, end-stage renal disease, and death^9^. Such disparities are likely driven by social determinants of health (SDOH)^10^, or the conditions in which people are born, live, work, and age^11^.

While SDOH generally describe community-level factors, health-related social needs (HRSN) refer to the specific social and economic needs that impact an individual’s health. Prior work suggests that HRSN, including access to care, education, gainful employment, and marital status, are strongly correlated with risk of hypertension and hypertension control^12^. Moreover, racial discrepancies in awareness and treatment of hypertension in the United States are prominent, with poorer awareness among Hispanic adults and lower likelihood of blood pressure control in Black vs. White adults^1,4^. Despite recognition of disparities in care delivery and hypertension outcomes^13^, there remains a persistent gap in the development and implementation of strategies to deliver more equitable cardiovascular care to minority females.

Even though HRSN may influence the development of hypertension, little is known about how cumulative social risk burden affects disease prevalence, particularly among underrepresented, minoritized women. The novel concept of cumulative social risk is adapted from genetics^14^, with a polygenic risk score representing an individual’s quantitative composite genetic risk for a disease^15^. Similarly, a “poly-social risk score” (PsRS) aggregates various social domains into a composite indicator of individual-level social risk for a given outcome^14^. A prior study assessing multiple domains of social needs, ranging from unemployment to psychological distress to food insecurity, found individuals with the highest PsRS had a nearly 4-fold higher prevalence of atherosclerotic cardiovascular disease (ASCVD) relative to those with the lowest risk scores after adjustment for conventional risk factors^7^. The authors also found that addition of social needs to traditional risk factors improved overall discrimination to detect ASCVD in a representative sample of US adults. However, the degree to which HRSN may help identify minority females with hypertension via a cumulative social disadvantage approach remains unknown. Furthermore, approaches to derive a PsRS for hypertension have not been previously studied specifically in this population.

To address these knowledge gaps, the aims of this study were (1) to test associations of cumulative and individual domains of HRSN with hypertension among Black and Hispanic women, and (2) to derive and validate a PsRS for hypertension among this population. We hypothesized that incorporating comprehensive poly-social risk into a predictive model would improve the identification of hypertension beyond conventional sociodemographic factors.

## METHODS

### Data Availability

All data and materials have been made publicly available at the Figshare data repository and can be accessed at https://figshare.com/articles/dataset/BUSY-BP_Dataset/30815915. The authors had full access to all the data in the study and assume responsibility for its integrity and data analysis.

### Study Cohort

Self-identified Black and Latina women aged 18-65 years who were receiving longitudinal care in the Mass General Brigham (MGB) healthcare system enrolled in the BUSY-BP study between February 2023 and April 2024. The MGB network provides care to 1.5 million patients across New England and includes two large academic medical centers, Massachusetts General Hospital and Brigham and Women’s Hospital, plus five other hospitals, affiliated practices, and outpatient centers. BUSY-BP participants were invited to participate in person, by telephone, or via the electronic medical record patient portal, in English and Spanish. The present analysis included all BUSY-BP participants who were not currently pregnant at the time of study enrollment. This study was approved by the MGB Institutional Review Board.

### Exposure: Health-Related Social Needs

On enrollment, participants completed the Centers for Medicare and Medicaid Services Accountable Health Communities Screening Tool (CMSST), adapted from previously validated instruments, to assess 13 domains of HRSN: housing instability, food insecurity, transportation problems, utility help needs, interpersonal safety, financial strain, employment difficulties, family and community support, education/training, physical activity, substance use, mental health, and disabilities. The questionnaire includes 26 questions (Supplemental Table 1), and each HRSN was examined individually as a candidate predictor. In addition, HRSN were aggregated into a cumulative domain score, with 1 point awarded for each domain with at least 1 positive response (range: 0-13).

### Covariates

On enrollment, participants completed demographic surveys including self-reported age, race/ethnicity, yearly household income, and highest level of education.

### Outcome: Hypertension

The primary outcome for this study was hypertension at enrollment, ascertained using a validated Electronic Health Record-based algorithm^16^ or the use of one or more antihypertensive medications for the indication of hypertension. Specifically, individuals were characterized as having hypertension if they either 1) had a qualifying International Classification of Diseases (ICD) code for hypertension (Supplemental Table 2) at two or more office visits, or 2) reported being diagnosed with hypertension and taking antihypertensive medications.

### Statistical Analyses

First, using the full cohort, we used logistic regression to examine associations of HRSN domains and individual HRSN with hypertension. We fit both unadjusted models and models adjusted for prespecified covariates selected as plausible confounders of this association (age, race/ethnicity, income, and education).

Next, we randomly divided the cohort into training (70%) and validation (30%) datasets. Least Absolute Shrinkage and Selection Operator (LASSO) regression with 10-fold cross-validation was used to derive a PsRS for hypertension in the training set. Race/ethnicity, income, education, and individual HRSN responses were included as potential covariates; age was included as a required coefficient in the final PsRS. The PsRS was tested in the validation set against a comparator model that included age, race/ethnicity, income, and education. Area under the receiver operator curve (AUC) in the validation set was compared across models using the Delong test.

Two-sided P<0.05 indicated statistical significance. Analysis was conducted using Stata 18.0.

## RESULTS

### Study cohort

A total of 1,302 women (mean [SD] age 40.1 [11.3] years, 70.4% Black, 44.3% Latina) were included (Table 1), and 29.3% of participants had hypertension. Across the entire cohort, the median overall domain score, with 1 point awarded per reported domain of HRSN, was 5 (interquartile range: 4-7, overall range 0-13) (Table 2). The most common HRSN domains among the cohort were family and community concerns (86%), mental health-related needs (82%), physical inactivity (76%), and financial strain (57%).

**Table 1:** Baseline Characteristics of Study Population.

| **Characteristic** | **Total Cohort**<br>**(N=1302)** | **Validation Set**<br>**(N=389)** | **Training Set**<br>**(N=913)** |
| --- | --- | --- | --- |
| Age (mean, SD) | 40.1 (11.3) | 39.9 (11.4) | 40.2 (11.3) |
| **Race** |  |  |  |
| White | 247 (19.0%) | 72 (18.5%) | 175 (19.2%) |
| Black or African American | 916 (70.4%) | 276 (71.0%) | 640 (70.1%) |
| American Indian or Alaska Native | 12 (0.9%) | 4 (1.0%) | 8 (0.9%) |
| Asian | 8 (0.6%) | 3 (0.8%) | 5 (0.5%) |
| Native Hawaiian or Pacific Islander | 4 (0.3%) | 3 (0.8%) | 1 (0.1%) |
| Multiracial or Biracial | 157 (12.1%) | 39 (10.0%) | 118 (12.9%) |
| Other | 210 (16.1%) | 60 (15.4%) | 150 (16.4%) |
| **Ethnicity** |  |  |  |
| Hispanic, Latino/Latina or of Spanish Origin | 577 (44.3%) | 170 (43.7%) | 407 (44.6%) |
| **Primary Language** |  |  |  |
| English | 1016 (78.0%) | 309 (79.4%) | 707 (77.4%) |
| Spanish | 243 (18.7%) | 67 (17.2%) | 176 (19.3%) |
| Arabic | 1 (0.1%) | 0 (0.0%) | 1 (0.1%) |
| Portuguese | 11 (0.8%) | 5 (1.3%) | 6 (0.7%) |
| Haitian Creole | 12 (0.9%) | 3 (0.8%) | 9 (1.0%) |
| French | 4 (0.3%) | 0 (0.0%) | 4 (0.4%) |
| Cape Verdean | 5 (0.4%) | 1 (0.3%) | 4 (0.4%) |
| Other | 10 (0.8%) | 4 (1.0%) | 6 (0.7%) |
| **Yearly Household Income** |  |  |  |
| Less than \$20,000 | 254 (19.5%) | 88 (22.6%) | 166 (18.2%) |
| \$20,000-\$49,000 | 304 (23.3%) | 81 (20.8%) | 223 (24.4%) |
| \$50,000-\$99,000 | 391 (30.0%) | 120 (30.8%) | 271 (29.7%) |
| \$100,000-\$150,000 | 172 (13.2%) | 48 (12.3%) | 124 (13.6%) |
| More than \$150,000 | 177 (13.6%) | 52 (13.4%) | 125 (13.7%) |
| No response | 4 (0.3%) | 0 (0.0%) | 4 (0.4%) |

| **Highest grade of school or college completed** |  |  |  |
| --- | --- | --- | --- |
| 8th grade or less | 11 (0.8%) | 6 (1.5%) | 5 (0.5%) |
| Some high school | 34 (2.6%) | 12 (3.1%) | 22 (2.4%) |
| High school graduate or GED | 249 (19.1%) | 73 (18.8%) | 176 (19.3%) |
| Technical degree or certificate | 119 (9.1%) | 35 (9.0%) | 84 (9.2%) |
| 1-3 years of college, including associates degree | 233 (17.9%) | 74 (19.0%) | 159 (17.4%) |
| 4 or more years of college, bachelor's degree | 362 (27.8%) | 110 (28.3%) | 252 (27.6%) |
| Graduate or professional school | 294 (22.6%) | 79 (20.3%) | 215 (23.5%) |
| **Employment Status** |  |  |  |
| Employed full-time (includes self-employed) | 810 (62.2%) | 220 (56.6%) | 590 (64.6%) |
| Employed part-time (includes self-employed) | 153 (11.8%) | 46 (11.8%) | 107 (11.7%) |
| Unemployed or laid-off | 75 (5.8%) | 23 (5.9%) | 52 (5.7%) |
| Unemployed due to disability | 81 (6.2%) | 31 (8.0%) | 50 (5.5%) |
| Full-time student | 58 (4.5%) | 23 (5.9%) | 35 (3.8%) |
| Homemaker | 60 (4.6%) | 22 (5.7%) | 38 (4.2%) |
| Retired | 22 (1.7%) | 8 (2.1%) | 14 (1.5%) |
| Other | 42 (3.2%) | 15 (3.9%) | 27 (3.0%) |
| No response | 1 (0.1%) | 1 (0.3%) | 0 (0.0%) |

**Table 2:**
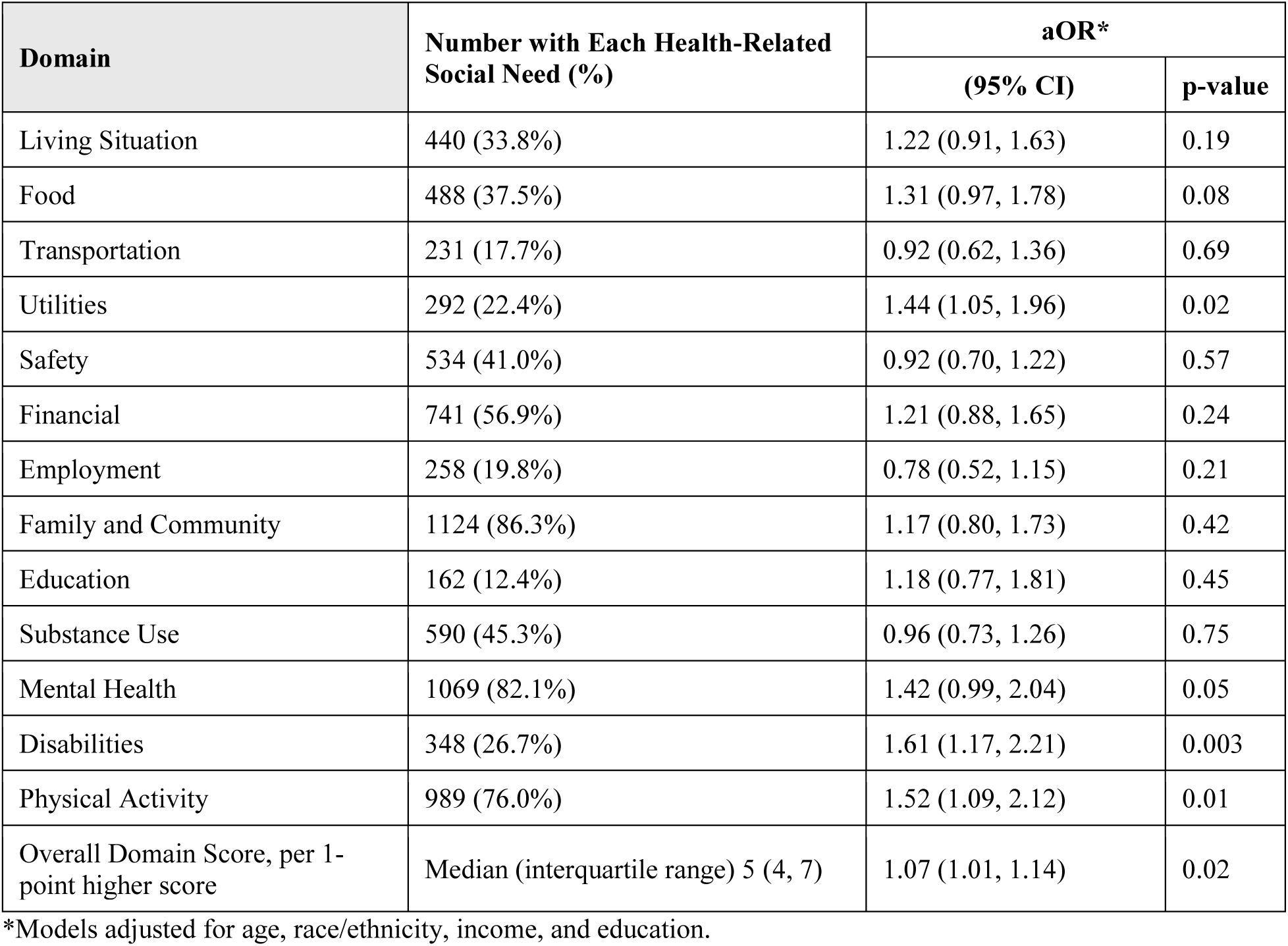
HRSN Domains and Adjusted Odds of Hypertension in the BUSY-BP Cohort (N=1,302).

| Domain | Number with Each Health-Related Social Need (%) | aOR* |  |
| --- | --- | --- | --- |
|  |  | (95% CI) | p-value |
| Living Situation | 440 (33.8%) | 1.22 (0.91, 1.63) | 0.19 |
| Food | 488 (37.5%) | 1.31 (0.97, 1.78) | 0.08 |
| Transportation | 231 (17.7%) | 0.92 (0.62, 1.36) | 0.69 |
| Utilities | 292 (22.4%) | 1.44 (1.05, 1.96) | 0.02 |
| Safety | 534 (41.0%) | 0.92 (0.70, 1.22) | 0.57 |
| Financial | 741 (56.9%) | 1.21 (0.88, 1.65) | 0.24 |
| Employment | 258 (19.8%) | 0.78 (0.52, 1.15) | 0.21 |
| Family and Community | 1124 (86.3%) | 1.17 (0.80, 1.73) | 0.42 |
| Education | 162 (12.4%) | 1.18 (0.77, 1.81) | 0.45 |
| Substance Use | 590 (45.3%) | 0.96 (0.73, 1.26) | 0.75 |
| Mental Health | 1069 (82.1%) | 1.42 (0.99, 2.04) | 0.05 |
| Disabilities | 348 (26.7%) | 1.61 (1.17, 2.21) | 0.003 |
| Physical Activity | 989 (76.0%) | 1.52 (1.09, 2.12) | 0.01 |
| Overall Domain Score, per 1-point higher score | Median (interquartile range) 5 (4, 7) | 1.07 (1.01, 1.14) | 0.02 |

\*Models adjusted for age, race/ethnicity, income, and education.

### Association of HRSN domains with hypertension

Each 1-point increase in the overall domain score was associated with an aOR of 1.07 (95% CI, 1.01–1.14, *P*=.02) for the presence of hypertension (Table 2). After adjustment for age, race/ethnicity, income, and education, screening positive for utility insecurity and having a disability were associated with 1.44 (95% CI 1.05-1.96, P=0.02) and 1.61 [95% CI 1.17-2.21, P=0.003] times higher odds of hypertension, respectively. Lack of physical activity in the prior month was also associated with 1.52-fold higher adjusted odds of hypertension (95% CI 1.09-2.12, P=0.01).

### Association of specific HRSN with hypertension

We next examined the 26 specific HRSN ascertained by the CMSST. The most frequently reported HRSN were occasional difficulties paying for food, housing and/or medical care (43.7%), feeling depressed several days a week (33.6%), physical inactivity (26.3%), and occasional loneliness (25.0%) (Supplemental Table I). Worries about food insecurity (adjusted odds ratio [aOR] 2.30, 95% CI 1.37-3.87, P= 0.002), lapse in utilities (aOR 1.44, 95% CI 1.04-1.96, P=0.02), social isolation (aOR 1.77, 95% CI 1.14-2.73, P=0.01) and difficulties concentrating, remembering, or making decisions because of a physical, mental, or an emotional condition (aOR 1.57, 95% CI 1.13-2.17, P=0.007) were associated with increased risk of hypertension (Table 3, Supplemental Table 3) after adjusting for age, race/ethnicity, income, and education. In contrast, moderate exercise between 1-4 days weekly (aOR 0.63, 95% CI 0.46-0.86, P=0.004) and 5-7 days weekly over the prior month (aOR 0.49, 95% CI 0.33-0.73, P<0.001) were associated with lower risk of hypertension (Table 3, Supplemental Table 3).

**Table 3:** Specific HRSN and Statistically Significant Adjusted Odds of HTN in the BUSY-BP Cohort (N=1,302).

| **Living Situation HRSN** | **aOR\* (95% CI)** | **P-value** |
| --- | --- | --- |
| Within the past 12 months, you worried that your food would run out before you got money to buy more. |  |  |
| Often true | 2.30 (1.37, 3.87) | 0.002 |
| Sometimes true | 1.31 (0.94, 1.82) | 0.11 |
| Never true | Ref |  |
| In the past 12 months has the electric, gas, oil, or water company threatened to shut off services in your home? |  |  |
| Never | Ref |  |
| Ever | 1.44 (1.04, 1.96) | 0.02 |
| **Family/Community Support HRSN** |  |  |
| How often do you feel lonely or isolated from those around you? |  |  |
| Never | Ref |  |
| Rarely | 1.46 (1.02, 2.07) | 0.03 |
| Sometimes | 1.14 (0.79, 1.63) | 0.48 |
| Often/Always | 1.77 (1.14, 2.73) | 0.01 |
| **Physical Activity HRSN** |  |  |
| In the last 30 days, other than the activities you did for work, on average, how many days per week did you engage in moderate exercise (like walking fast, running, jogging, dancing, swimming, biking, or other similar activities)? |  |  |
| 0 days | Ref |  |
| 1-4 | 0.63 (0.46, 0.86) | 0.004 |
| 5-7 | 0.49 (0.33, 0.73) | <0.001 |
| **Disabilities HRSN** |  |  |
| Because of a physical, mental, or emotional condition, do you have serious difficulty concentrating, remembering, or making decisions? (5 years old or older) | 1.57 (1.13, 2.17) | 0.007 |

\*Models adjusted for age, race/ethnicity, income, and education.

### Hypertension prediction

Using sociodemographic characteristics and detailed HRSN responses, we next trained a PsRS for hypertension in the training set and compared its performance to that of the sociodemographic model (age, income, race/ethnicity, education) in the validation set. In the training set, the per-standard deviation odds of hypertension were 2.91 (95% CI 2.43-3.47, P<0.001) for the sociodemographic model and 3.30 (95% CI 2.73-3.99, P<0.001) for the PsRS. The PsRS improved discrimination for HTN beyond conventional demographic indicators in the training set (PsRS: AUC 0.79 [95% CI 0.75-0.82] vs. sociodemographic model: AUC 0.77 [95% CI 0.73-0.80], P=0.008) (Figure 1). However, the PsRS was not superior to the sociodemographic model in the validation set (PsRS: AUC 0.76 [95% CI 0.71-0.81] vs. sociodemographic model: AUC 0.80 [95% CI 0.75-0.85], P=0.005) (Figure 1).

**Figure 1.**
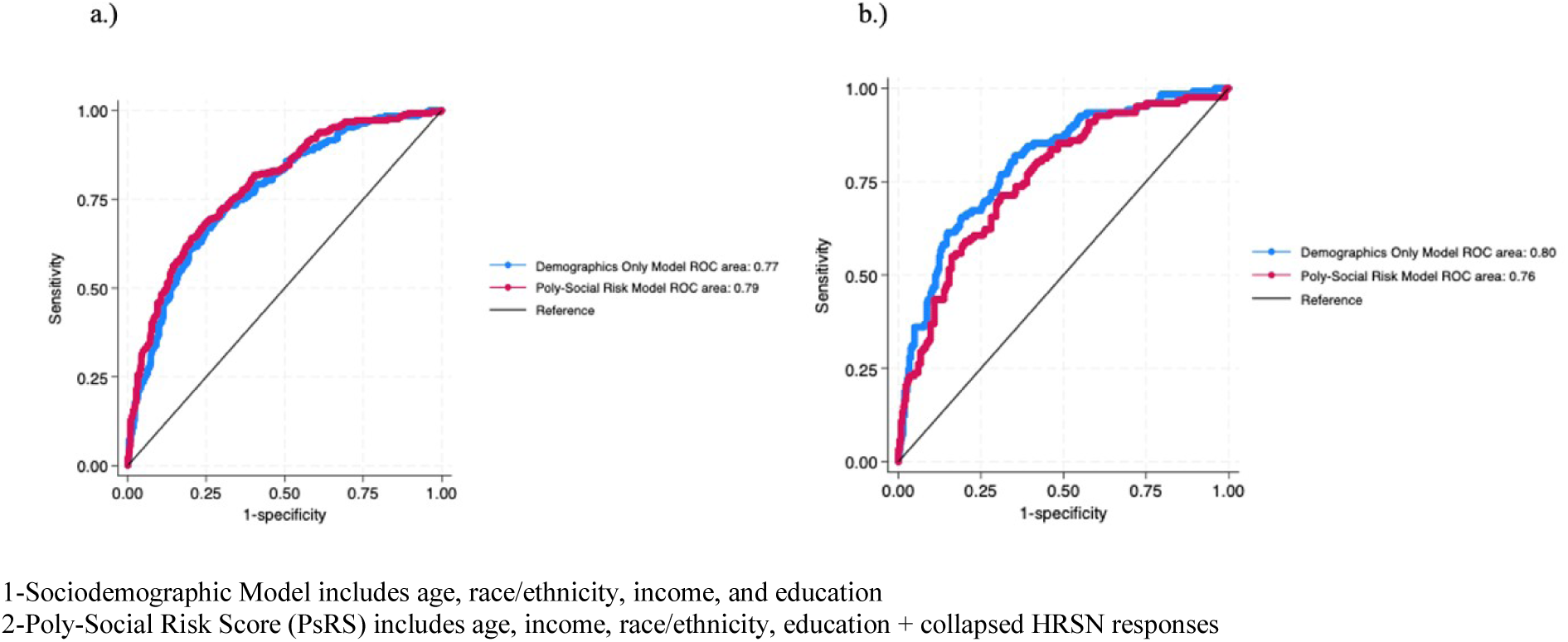
Receiver Operating Curves (ROC) Comparing the Sociodemographic Model^1^ and Poly-Social Risk Score^2^ (PsRS) in the Training Set (a) and Validation Set (b) 1-Sociodemographic Model includes age, race/ethnicity, income, and education 2-Poly-Social Risk Score (PsRS) includes age, income, race/ethnicity, education + collapsed HRSN responses

## DISCUSSION

In this study, we investigated the association of cumulative social disadvantage, as well as individual social needs, with hypertension prevalence among Black and Latina females. Our findings suggested that a higher overall burden of HRSN was associated with increased risk of hypertension, independent of age, income, race, and education level. The broad social domains of physical inactivity, having a disability, and utility insecurity were associated with hypertension. With respect to specific HRSN, worries related to food insecurity were associated with the greatest relative risk, more than two-fold odds of hypertension, followed by social isolation, poor concentration or decision making, and lapse in utilities. In contrast, physical activity was the only significant protective social factor in our cohort, with routine weekly exercise associated with nearly a 40% reduction in the odds of hypertension. Despite associations between specific social needs and hypertension in our study population, incorporating granular HRSN into a predictive model did not improve hypertension detection beyond conventional sociodemographic indices.

Our results permit several conclusions. First, greater cumulative social disadvantage is associated with modestly increased risk of hypertension among Black and Latina women. These findings mirror those from a nationally representative sample of U.S. adults (mean [SD] age 62.0 [10.2] years), which revealed higher social disadvantage among non-Hispanic Black and Hispanic adults was also associated with mildly higher odds of hypertension (aOR 1.17 95% CI [0.99–1.38]) relative to non-Hispanic White adults^17^. Additional research has also shown a dose–response association between number of adverse SDOH and hypertension prevalence in U.S. adults^18^. This trend has been postulated to arise from the complex interplay of behavioral and clinical risk factors, chronic stress exposure, and pro-inflammatory dysregulation^19,20^, further amplified by greater social deprivation. The exact mechanisms by which cumulative social disadvantage confers cardiovascular risk among minoritized females, coupled with assessment of targeted interventions to address unmet social needs, warrant future investigation.

Second, specific HRSN may be particularly relevant to hypertension risk. Our results redemonstrate the well-established relationships between food insecurity^21,22^ and physical inactivity^23–25^ and hypertension. However, our study also identifies utility instability, social isolation, having a disability, and poor concentration as risk factors for hypertension among female minorities. These findings provide insights into potentially underrecognized psychosocial needs that may confer increased cardiovascular vulnerability among Black and Latina women and may indicate subgroups of women who benefit from primordial prevention interventions to prevent the development of hypertension.

Third, despite the associations between specific HRSN and hypertension prevalence, granular HRSN may have limited predictive utility for identifying Black and Latina women at highest risk of hypertension beyond more conventional sociodemographic indices. In contrast to our initial hypothesis, addition of HRSN to traditional sociodemographic risk factors did not improve overall prediction of hypertension. It is plausible social factors that impart high risk for hypertension were not captured by our model, justifying further investigation of potentially more nuanced health-related needs among this population. Given the associations we observed between certain social factors and hypertension, consideration of HRSN may be more useful for informing targeted interventions to minimize risk, rather than predict disease.

Finally, although HRSN did not meaningfully improve hypertension discrimination beyond conventional sociodemographic factors, screening for social needs may still help identify actionable barriers to cardiovascular health. Current ACC/AHA guidelines endorse routine SDOH assessment^27^, and prior intervention studies targeting food insecurity^28^ and physical inactivity^29^ have demonstrated improvements in cardiometabolic outcomes among Black and Latina women. Future work should evaluate whether HRSN-guided interventions can improve hypertension prevention and control in minoritized women.

### Strengths and limitations

Our study has certain strengths, including a focus on Black and Latina women, who are underrepresented in CVD research, yet overrepresented in CVD burden. Furthermore, HRSN were evaluated comprehensively via the CMSST, providing more granular detail on factors that may predispose minorities to hypertension.

Our study also has limitations. First, its focus on individuals receiving care in an integrated healthcare system in eastern Massachusetts, where insurance rates are high, may suggest our results are not generalizable to minority females in other areas of the country. Second, HRSN were self-reported, which may introduce recall bias and misclassification. Third, our study is cross-sectional in nature, and thus causality between HRSN and hypertension cannot be assessed. Finally, our study was not designed to assess the relationship between HRSN and severity of hypertension, or risk of hypertension-related complications over time. Future studies are needed to evaluate whether greater burden of HRSN is associated with more severe hypertension, and whether HRSN may predict risk of hypertension-related adverse outcomes longitudinally.

## Conclusions

Our findings reveal key hypertension-relevant HRSN among a unique cohort of Black and Latina individuals and indicate that greater cumulative burden of HRSN is associated with heightened odds of hypertension among female minorities. Screening for HRSN in healthcare settings may serve as an opportunity to identify and address the social needs that place minoritized individuals at greatest risk for cardiovascular disease. These findings may also inform future investigation of targeted interventions to optimize cardiovascular health and mitigate racial and ethnic disparities among females with hypertension.

## Non-standard Abbreviations and Acronyms

CVD: Cardiovascular disease
SDOH: social determinants of health
HRSN: health-related social needs
PsRS: poly-social risk score
ASCVD: atherosclerotic cardiovascular disease
MGB: Massachusetts General Brigham healthcare network
CMSST: Centers for Medicare and Medicaid Services Accountable Health Communities Screening Tool
ICD: International Classification of Diseases
LASSO: Least Absolute Shrinkage and Selection Operator Regression
AUC: Area under the receiver operator curve

## SUPPLEMENTAL MATERIAL

**Supplemental Table I:**
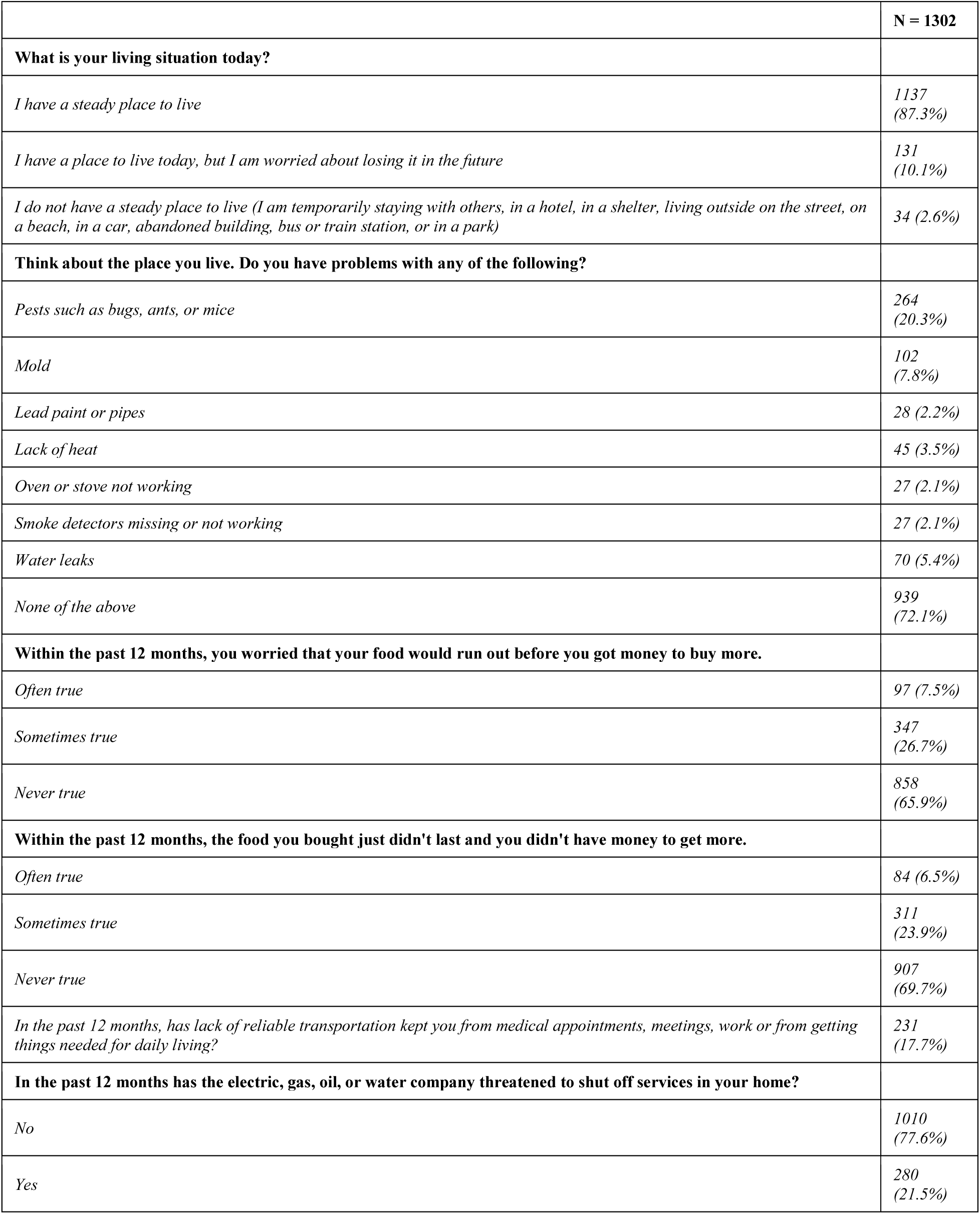
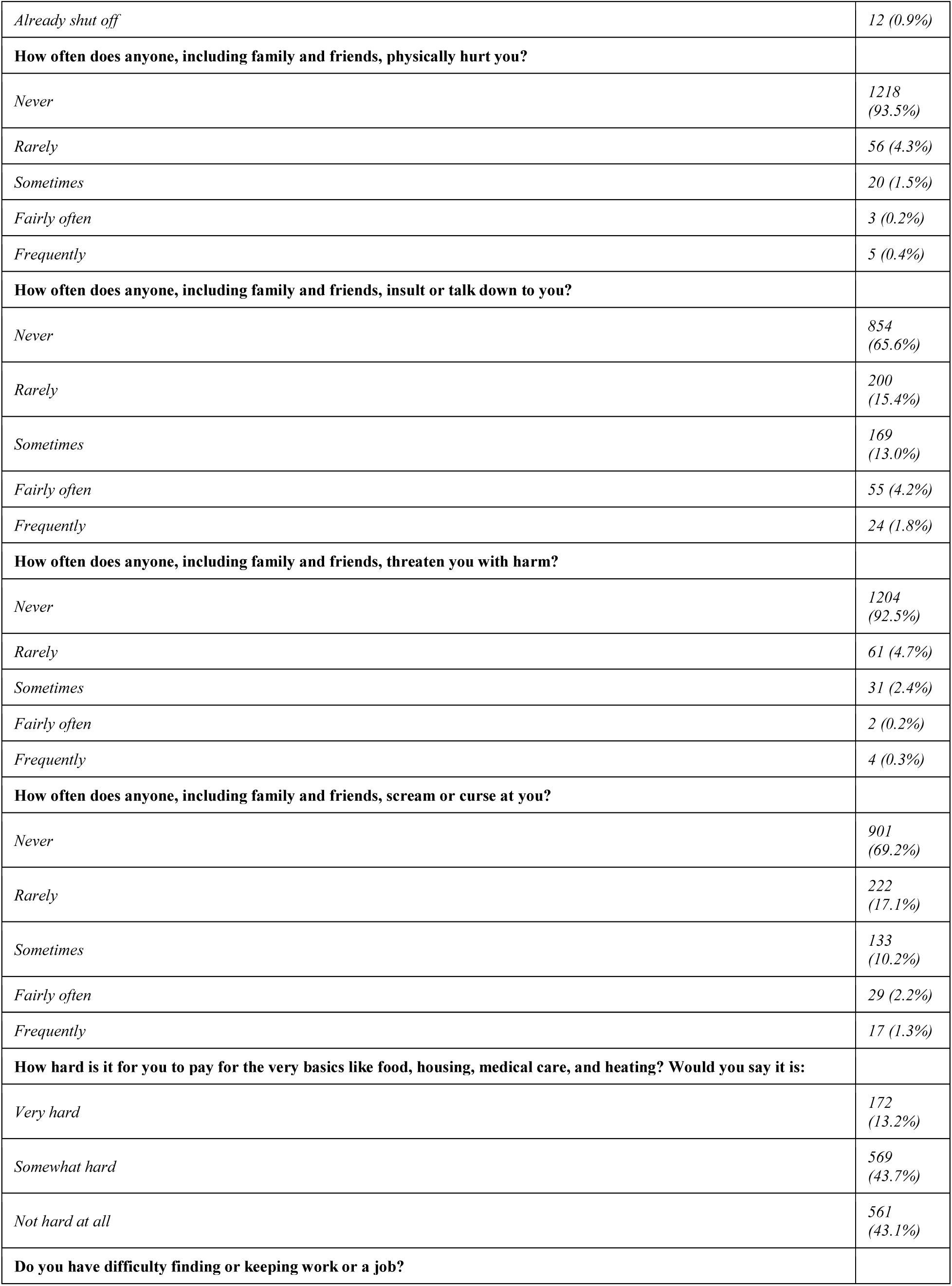
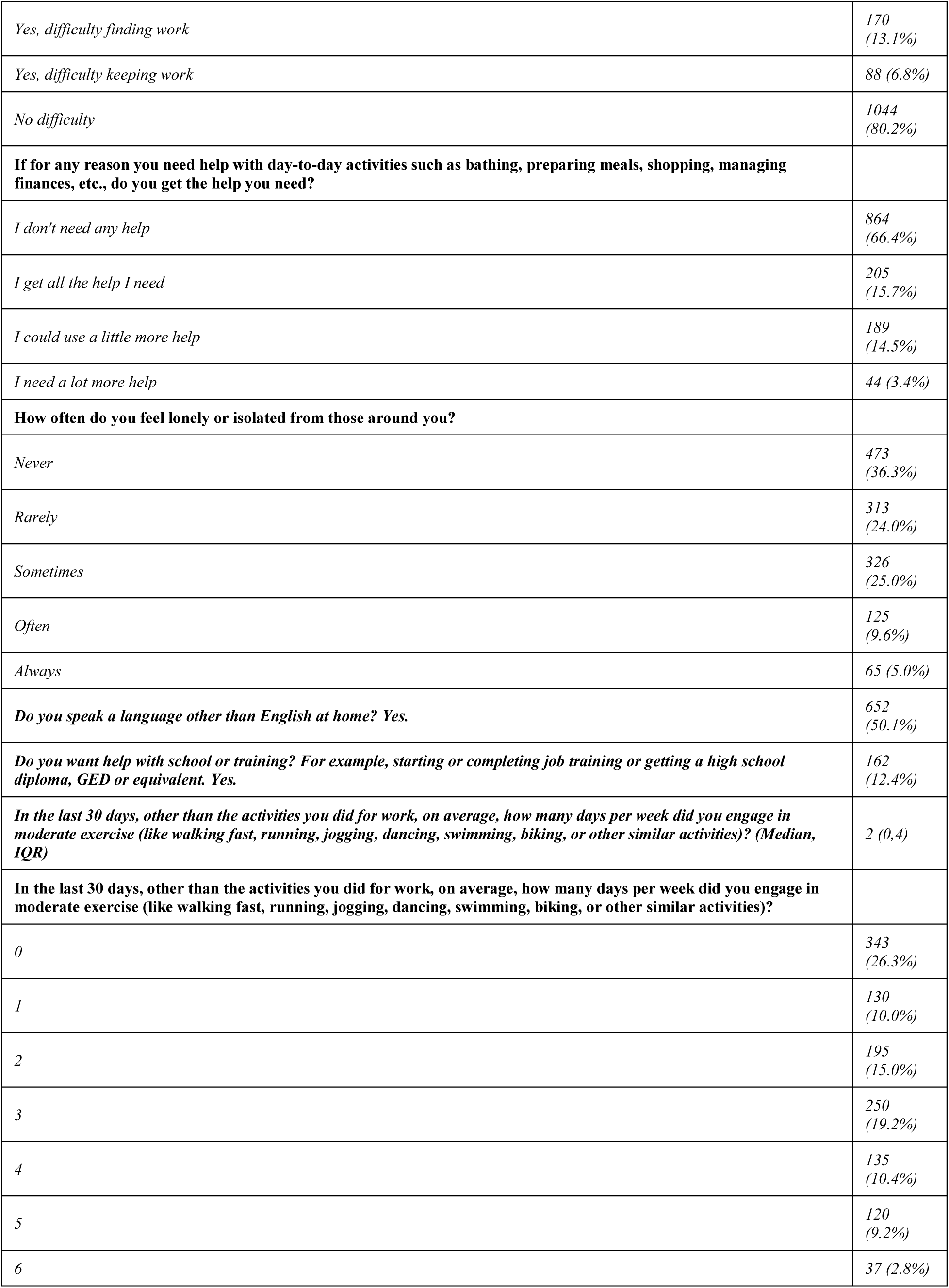
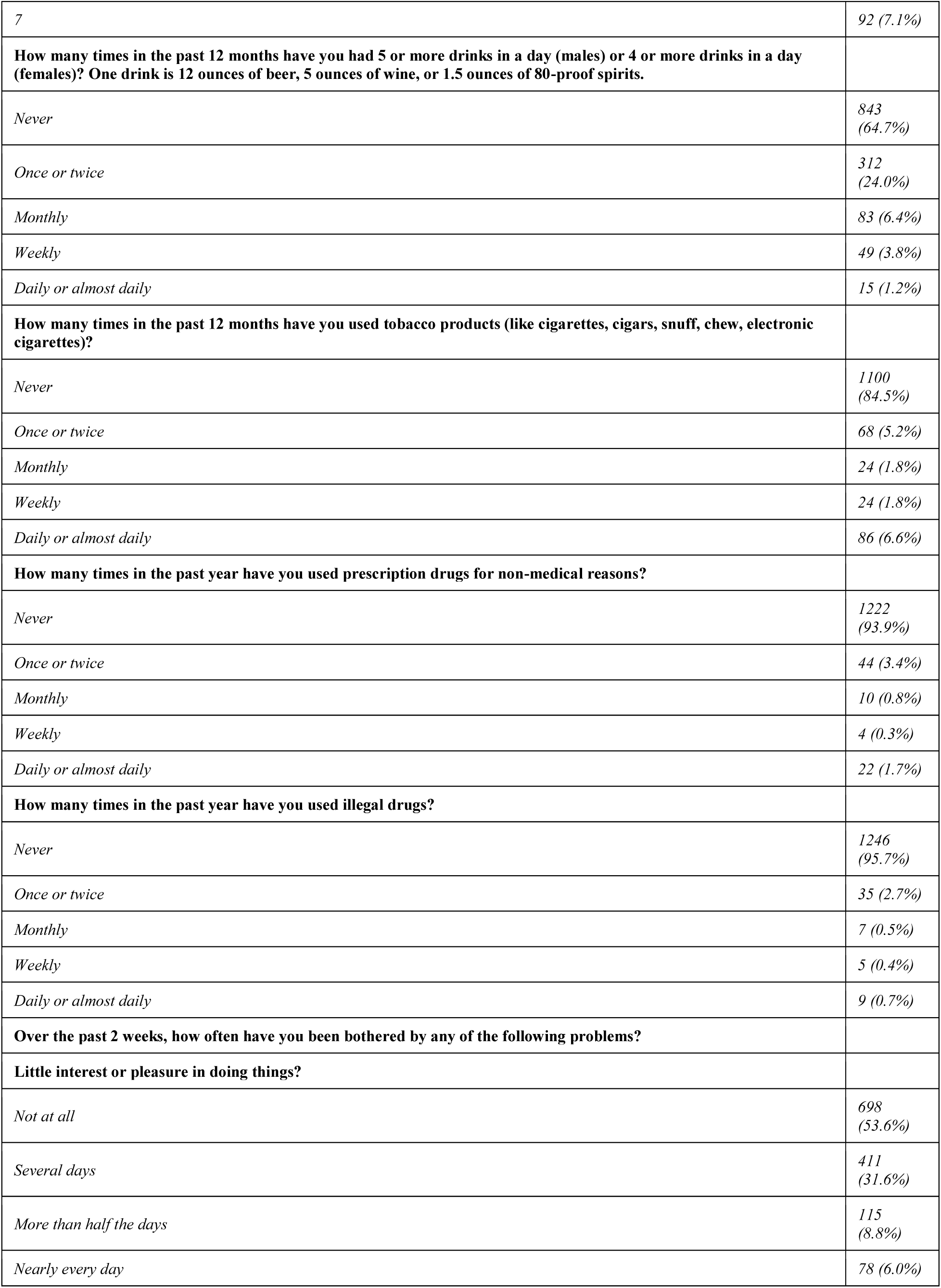
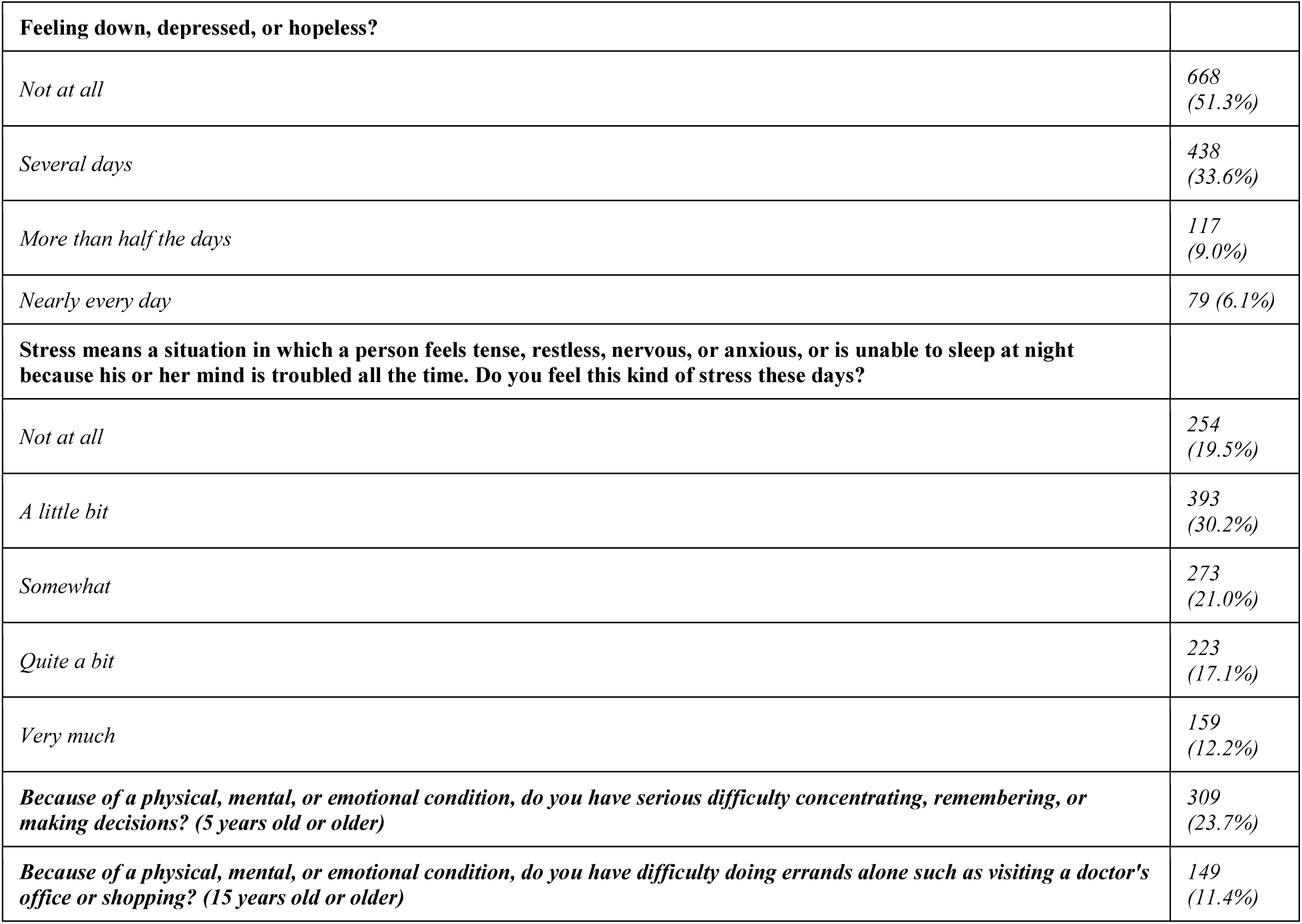
HRSN Questionnaire Responses Among the BUSY-BP Cohort.

**Supplemental Table II:** ICD Codes Used to Identify Hypertension.

| **Code** | **Description** |
| --- | --- |
| **ICD-9 Codes (use before 10/1/15)** |  |
| 997.91 | Hypertension |
| 401.0 | Malignant essential hypertension |
| 401.1 | Benign essential hypertension |
| 401.9 | Unspecified essential hypertension |
| 402.01 | Malignant hypertensive heart disease with congestive heart failure |
| 402.00 | Malignant hypertensive heart disease without congestive heart failure |
| 402.91 | Unspecified hypertensive heart disease with congestive heart failure |
| 402.90 | Unspecified hypertensive heart disease without congestive heart failure |
| 403.01 | Malignant hypertensive renal disease with renal failure |
| 403.00 | Malignant hypertensive renal disease without mention of renal failure |
| 404.01 | Malignant hypertensive heart and renal disease with congestive heart failure |
| 404.03 | Malignant hypertensive heart and renal disease with congestive heart failure and renal failure |
| 404.91 | Unspecified hypertensive heart and renal disease with congestive heart failure |
| 404.93 | Unspecified hypertensive heart and renal disease with congestive heart failure and renal failure |
| 404.92 | Unspecified hypertensive heart and renal disease with renal failure |
| 404.11 | Benign hypertensive heart and renal disease with congestive heart failure |
| 404.13 | Benign hypertensive heart and renal disease with congestive heart failure and renal failure |
| 404.12 | Benign hypertensive heart and renal disease with renal failure |
| 404.10 | Benign hypertensive heart and renal disease without mention of congestive heart failure or renal failure |
| 402.11 | Benign hypertensive heart disease with congestive heart failure |
| 402.10 | Benign hypertensive heart disease without congestive heart failure |
| 403.11 | Benign hypertensive renal disease with renal failure |
| 403.10 | Benign hypertensive renal disease without mention of renal failure |
| **ICD-10 Codes (after 10/1/15)** |  |
| I10 | Essential (primary) hypertension |
| I11.0 | Hypertensive heart disease with heart failure |
| I11.9 | Hypertensive heart disease without heart failure |
| I13.0 | Hypertensive heart and chronic kidney disease with heart failure and stage 1 through stage 4 chronic kidney disease, or unspecified chronic kidney disease |
| I13.10 | Hypertensive heart and chronic kidney disease without heart failure with stage 1 through stage 4 chronic kidney disease, or unspecified chronic kidney disease |
| I13.11 | Hypertensive heart and chronic kidney disease without heart failure with stage 5 chronic kidney disease, or end stage renal disease |
| I13.2 | Hypertensive heart and chronic kidney disease with heart failure and with stage 5 chronic kidney disease, or end stage renal disease |
| I15.9 | Secondary hypertension, unspecified |

**Supplemental Table III:** All HRSN and Adjusted Odds of HTN Among the BUSY-BP Cohort (N=1,302).

| HRSN Factors | aOR* (95% CI) | P-value |
| --- | --- | --- |
| **Living Situation Questions** |  |  |
| **What is your living situation today?** |  |  |
| I have a steady place to live | Ref |  |
| I do not have a steady place to live | 1.13 (0.75, 1.71) | 0.57 |
| **Think about the place you live. Do you have problems with any of the following (pests, mold, lead paint/pipes, oven/stove not working, smoke detectors, water leaks)?** |  |  |
| No | Ref |  |
| Yes | 1.25 (0.92, 1.70) | 0.14 |
| **Within the past 12 months, you worried that your food would run out before you got money to buy more.** |  |  |
| Often true | 2.30 (1.37, 3.87) | 0.002 |
| Sometimes true | 1.31 (0.94, 1.82) | 0.11 |
| Never true | Ref |  |
| **Within the past 12 months, the food you bought just didn't last and you didn't have money to get more.** |  |  |
| Often true | 1.53 (0.88, 2.69) | 0.13 |
| Sometimes true | 1.40 (0.99, 1.96) | 0.05 |
| Never true | Ref |  |
| In the past 12 months, has lack of reliable transportation kept you from medical appointments, meetings, work or from getting things needed for daily living? | 0.92 (0.62, 1.36) | 0.69 |
| **In the past 12 months has the electric, gas, oil, or water company threatened to shut off services in your home?** |  |  |
| Never | Ref |  |
| Ever | 1.44 (1.04, 1.96) | 0.02 |
| **Safety** |  |  |
| **How often does anyone, including family and friends, physically hurt you?** |  |  |
| Never | Ref |  |
| Ever | 1.18 (0.70, 2.02) | 0.54 |
| **How often does anyone, including family and friends, insult or talk down to you?** |  |  |
| Never | Ref |  |
| Rarely | 0.94 (0.64, 1.38) | 0.75 |
| Sometimes | 1.14 (0.75, 1.72) | 0.54 |
| Fairly often/frequently | 0.85 (0.48, 1.52) | 0.59 |
| **How often does anyone, including family and friends, threaten you with harm?** |  |  |
| Never | Ref |  |
| Ever | 1.11 (0.66, 1.85) | 0.70 |
| **How often does anyone, including family and friends, scream or curse at you?** |  |  |
| Never | Ref |  |
| Rarely | 0.85 (0.58, 1.25) | 0.41 |
| Sometimes/fairly often/frequently | 1.10 (0.74, 1.63) | 0.63 |
| **Financial Strain, Employment, Family, Community Support, Education Questions** |  |  |
| **How hard is it for you to pay for the very basics like food, housing, medical care, and heating? Would you say it is:** |  |  |
| Very hard | 1.26 (0.79, 2.02) | 0.34 |
| Somewhat hard | 1.19 (0.86, 1.65) | 0.28 |
| Not hard at all | Ref |  |
| **Do you have difficulty finding or keeping work or a job?** |  |  |
| Yes, difficulty finding work | 0.82 (0.52, 1.30) | 0.40 |
| Yes, difficulty keeping work | 0.70 (0.39, 1.25) | 0.23 |
| No difficulty | Ref |  |
| **If for any reason you need help with day-to-day activities such as bathing, preparing meals, shopping, managing finances, etc., do you get the help you need?** |  |  |
| I don't need any help | Ref |  |
| I get all the help I need | 1.16 (0.80, 1.69) | 0.43 |
| I could use more help | 1.31 (0.91, 1.88) | 0.14 |
| **How often do you feel lonely or isolated from those around you?** |  |  |
| Never | Ref |  |
| Rarely | 1.46 (1.02, 2.07) | 0.03 |
| Sometimes | 1.14 (0.79, 1.63) | 0.48 |
| Often/Always | 1.77 (1.14, 2.73) | 0.01 |
| Do you speak a language other than English at home? | 0.86 (0.62, 1.19) | 0.37 |
| Do you want help with school or training? For example, starting or completing job training or getting a high school diploma, GED or equivalent. | 1.18 (0.77, 1.81) | 0.45 |
| **Physical Activity Questions** |  |  |
| **In the last 30 days, other than the activities you did for work, on average, how many days per week did you engage in moderate exercise (like walking fast, running, jogging, dancing, swimming, biking, or other similar activities)?** |  |  |
| 0 days | Ref |  |
| 1-4 | 0.63 (0.46, 0.86) | 0.004 |
| 5-7 | 0.49 (0.33, 0.73) | <0.001 |
| **Substance Use, Mental Health, Disabilities Questions** |  |  |
| **How many times in the past 12 months have you had 5 or more drinks in a day (males) or 4 or more drinks in a day (females)? One drink is 12 ounces of beer, 5 ounces of wine, or 1.5 ounces of 80-proof spirits.** |  |  |
| Never | Ref |  |
| Ever | 1.08 (0.81, 1.45) | 0.59 |
| **How many times in the past 12 months have you used tobacco products (like cigarettes, cigars, snuff, chew, electronic cigarettes)?** |  |  |
| Never | Ref |  |
| Occasional | 0.83 (0.50, 1.40) | 0.49 |
| Daily/Almost Daily | 1.12 (0.66, 1.91) | 0.67 |
| **How many times in the past year have you used prescription drugs for non-medical reasons?** |  |  |
| Never | Ref |  |
| Ever | 0.75 (0.43, 1.36) | 0.36 |
| **How many times in the past year have you used illegal drugs?** |  |  |
| Never | Ref |  |
| Ever | 0.91 (0.45, 1.85) | 0.80 |
| **Over the past 2 weeks, how often have you been bothered by any of the following problems?** |  |  |
| **Little interest or pleasure in doing things?** |  |  |
| Not at all | Ref |  |
| Several days | 1.28 (0.94, 1.75) | 0.11 |
| More than half the days | 1.56 (0.95, 2.56) | 0.08 |
| Nearly every day | 1.37 (0.76, 2.47) | 0.30 |
| **Feeling down, depressed, or hopeless?** |  |  |
| Not at all | Ref |  |
| Several days | 1.16 (0.86, 1.56) | 0.32 |
| More than half the days | 1.64 (1.00, 2.70) | 0.05 |
| Nearly every day | 1.16 (0.65, 2.10) | 0.63 |
| **Stress means a situation in which a person feels tense, restless, nervous, or anxious, or is unable to sleep at night because his or her mind is troubled all the time. Do you feel this kind of stress these days?** |  |  |
| Not at all | Ref |  |
| A little bit | 1.22 (0.82, 1.80) | 0.33 |
| Somewhat | 1.27 (0.83, 1.94) | 0.27 |
| Quite a bit | 1.59 (1.01, 2.50) | 0.05 |
| Very much | 1.54 (0.92, 2.56) | 0.09 |
| **Because of a physical, mental, or emotional condition, do you have serious difficulty concentrating, remembering, or making decisions? (5 years old or older)** | 1.57 (1.13, 2.17) | 0.007 |
| **Because of a physical, mental, or emotional condition, do you have difficulty doing errands alone such as visiting a doctor's office or shopping? (15 years old or older)** | 1.75 (1.14, 2.71) | 0.01 |

\*Models adjusted for age, race, income, and education

## PERSPECTIVES

Among Black and Latina women, greater cumulative burden of health-related social needs was associated with higher odds of hypertension, independent of age, race/ethnicity, income, and education. Specific needs—including food insecurity, utility insecurity, social isolation, and cognitive difficulties—were particularly strongly associated with hypertension, while regular physical activity was associated with lower odds. Although incorporation of detailed health-related social needs did not improve hypertension identification beyond conventional sociodemographic factors, screening for these needs may help identify actionable barriers to cardiovascular health. Future studies should evaluate whether interventions targeting unmet social needs can improve hypertension prevention and control and help reduce persistent cardiovascular health disparities among minoritized women.

## NOVELTY AND RELEVANCE

### What is New?

Black and Latina women are disproportionately affected by hypertension in the United States, which is in large part driven by social disadvantage. It remains unclear which social factors impart greatest risk, and whether cumulative burden of social needs can improve detection of hypertension in this population.

### What is Relevant?

A greater aggregate burden of health-related social needs was associated with higher odds of hypertension in Black and Latina women. Among specific HRSN, food and utility insecurity, social isolation, and poor concentration were each associated with increased risk of hypertension. However, poly-social risk did not improve prediction of hypertension beyond conventional sociodemographic measures.

### Clinical/Pathophysiological Implications?

Further investigation of targeted interventions guided by HRSN screenings may help mitigate inequities in hypertension outcomes among racially diverse females.

